# Utility of gut-brain electrophysiological coupling in predicting L-Dopa induced dyskinesia in Parkinson’s Disease

**DOI:** 10.1101/2024.12.04.24318228

**Authors:** Sanket Houde, Mansimran Kaur, Hari Prakash Tiwari, Nandini Priyanka B, Rathore BP, Pragathi P. Balasubramani

## Abstract

In this study, we focus on Levodopa induced dyskinesia (LID) condition in Parkinson’s Disease (PD) and investigate the mechanistic role of gut-brain coupling in explaining the dyskinesia severity. Earlier studies have provided evidences for abnormal dynamics in the cortico-basal ganglia loops and also in the gut functioning, for explaining LID. However to our knowledge, assessing gut-brain coupling isn’t a standard practice for development of the treatment strategy in LID patients for understanding the odds of dyskinesia onset and progression. In this study, we use scalable data acquisition technologies such as electroencephalography (EEG) and electrogastrography (EGG) for investigating the gut-brain coupling, and for the first time assess its utility to inform about dyskinesia severity in PD patients. We collected data from N=67 subjects (healthy = 26) in middle to old age adulthood and acquire their gut-brain coupling data during various cognitive engagement tasks, using simultaneous EEG and EGG recording setup. Some of our results include that gut-brain coupling can predict the severity of dyskinesia in PD during interoception, especially in hyperventilating and eyes closed resting state paradigms. Second, specific frequencies of gut coupling activity are more sensitive to explaining motor complications. Third, the gut activity differentially couples with different brain regions to explain the symptom severity. The most salient features of our model were the normogastric gut coupling with the temporo-occipital brain region, normogastric gut coupling with the frontal region, and the tachygastric gut coupling with the centro-parietal region. Furthermore of translational significance, the latter two features also significantly interacted with cardiac measures, and the model was able to predict sensitive heart rate variability levels for reducing the symptom severity. Altogether, our study paves way for utitlizing gut-brain coupling as a clinical measure for strategizing interventions in PD.

## Main

Parkinson’s disease (PD) is one of the most prevalent neurodegenerative disorders, suggested to be increasingly debilitating over time in humans, with symptomatic manifestations including motor disabilities such as rigidity, tremor, postural instability, cognitive impairments, mental illness, sleep disturbances (Dorsey et al., 2018; Jellinger, 2019). In India, the average onset of PD is as much as a decade younger than many other countries, at 51.03 ± 11.32 years. Some form of dopamine precursor such as levodopa (L-dopa) is presently the widely used medication for treating PD. With the advancement of the disease, L-dopa treatment is associated with the occurrence of involuntary movements, a condition called as L-dopa-induced dyskinesia (LID). Studies suggest that more than 80% of PD patients have the risk of developing LID, and about 30% of them may develop LID within just 3 years after L-dopa treatment (Ahlskog & Muenter, 2001; Fahn et al., 2004) due to some abnormal neural dynamics triggered by the intervention (Kim et al., 2020). Given the risk of PD onset in relatively younger population in India, the risk of dyskinesia in young patients is arguably worrisome (*Early Onset Parkinsonism (EOPD) in India vs. Western Populations*, n.d.).

On the other hand, the role of gut in Parkinson’s have been recognized strongly in the recent times: Approximately 60–80% of patients with PD experience gastrointestinal (GI) symptoms as soon as by 4 years post-PD diagnosis. Gastric emptying through the intestines is considered as the rate-limiting step in levodopa absorption, and prolonged exposure of levodopa in the stomach and small intestine increases the chances of its breakdown by enzymes, and decreasing its availability for absorption altogether causing impairments. This delayed gastric emptying in the patients observed through reduced stomach motility may be referred by the condition called gastroparesis in PD (Heimrich et al., 2019; Menozzi et al., 2021; Pfeiffer et al., 2020).

Recent studies including our own (Balasubramani et al., 2022) have shown that electrogastrography is a sensitive tool to observe the changes in gastric motility (EGG). Much like the electrical rhythms of the brain and heart, the gastroenterological (GI) tract has electrical activity governed by the enteric nervous system. The electrical wave propagating along the serosal surface of the stomach, the gastric slow wave, oscillates at approximately 3 cycles per minute (0.05 Hz) and coordinates the smooth muscle contractions of peristalsis during digestion. The waves generated at the stomach’s surface propagate to the skin via volume conduction, and these voltages can be measured with cutaneous electrodes. EGG as a methodology represents a scalable and non-invasive method to study brain-viscera interactions in psychophysiology and neurophysiology (Azzalini et al., 2019; Gharibans et al., 2017, 2019; Rebollo et al., 2018, 2021; Wolpert et al., 2019).

These advances with EGG create a unique opportunity to investigate the association between visceral bodily signals with neurophysiological brain measures studied with electroencephalography (EEG). The EGG measures will aid in developing a quantified understanding of the subjective gut state, contextualizing and assisting in predicting the behavior, cognition and mental health, and even relate to EEG-identified neurophysiological underpinnings. Recent studies, using high resolution magnetoencephalography (MEG) and EGG have shown that during resting state, rhythmic gut physiological signals are linked with cortical brain oscillations. Specifically, in a 2016 study, Richter et al. found significant phase-amplitude coupling between the infra-slow gastric phase and the amplitude of the cortical alpha rhythm (Richter et al., 2017). This coupling was localized to the right anterior insula and bilaterally to occipito-parietal regions and was noted to be due to ascending influences from the stomach to these brain regions. In 2018, Rebollo et al. evaluated gut-brain coupling by simultaneously recording fMRI and EGG in healthy volunteers (Rebollo et al., 2018). They discovered an elaborate gastric network cutting across classical resting-state networks with partial overlap with autonomic regulation areas.

In a nutshell, gut-brain coupling has not been investigated for its role during various cognitive brain engagements for predicting symptom severity in PD. In this study, we specifically investigate whether gut-brain coupling can inform about involuntary movements in dyskinesia observed due to L-dopa intake in PD. We hypothesize that electrogastrogram features can be a sensitive tool to measure the gut-brain coupling along with brain electrophysiology, and the coupling can predict the dyskinesia severity in PD. We also hypothesize that the gut signals feed into the brain for mediating the motor complications, and that the whole body cognition can help the prognosis and treatment plan for LID.

### Study cohort and differences in demographics and physiology

We acquired simultaneous electroencephalography (EEG) and electrogastrography (EGG) from two groups of participants, one suffering from Parkinson’s Disease with dyskinesia (59 ± 10.5 years, 69% males, UPDRS score for dyskinesia = 2.9 ± 2.08) and another healthy controls (70 ± 7.6 years, 50% males, UPDRS score for dyskinesia = 0.08 ± 0.41). The CONSORT chart for our study is presented in Figure 1. We used 19 channel EEG, and 7 channel EGG electrodes for our recording. Figure 2a presents a schematic of our setup. For simplicity, we focus on 3 broad brain regions for our analysis-frontal, centro-parietal, temporo-occipital, for investigating our hypothesis; the channels averaged for each of the regional categories are again shown in Figure 2a (also refer to Methods). Furthermore in EEG, we focus on beta frequency band of the data spectrum, for its significance in representing motor functionalities and control (Bai et al., 2007; Jenkinson & Brown, 2011; Ricci et al., 2019). Not surprisingly, the power topography (shown in Figure 2b) shows larger power around the motor strip of the cortex in the central brain. We observe the gut power spectral density had an overall peak at frequency in the *bradygastric* range at 0.013 Hz with power 5.8e7 μV^2^ across our cohorts. Generally, the healthy controls had EGG power much larger than PD in pre-intervention, and the post-pre intervention difference, referred as delta-condition in our text was significantly different between PD and healthy controls. In particular, the differences were significant in the *normogastric* range, with 7.1e4 μV^2^ in PD patients and it was −1.9e5 in healthy controls in the delta condition (Figure 2c, Table 1, p=0.01). The EEG-EGG coupling strength as computed using Phase Amplitude Coupling between the broadband EGG and beta signals of all EEG electrodes (see Methods, (Özkurt & Schnitzler, 2011), had a topographical profile differed to that of the brain power (Figure 2b).

**Figure 1:**
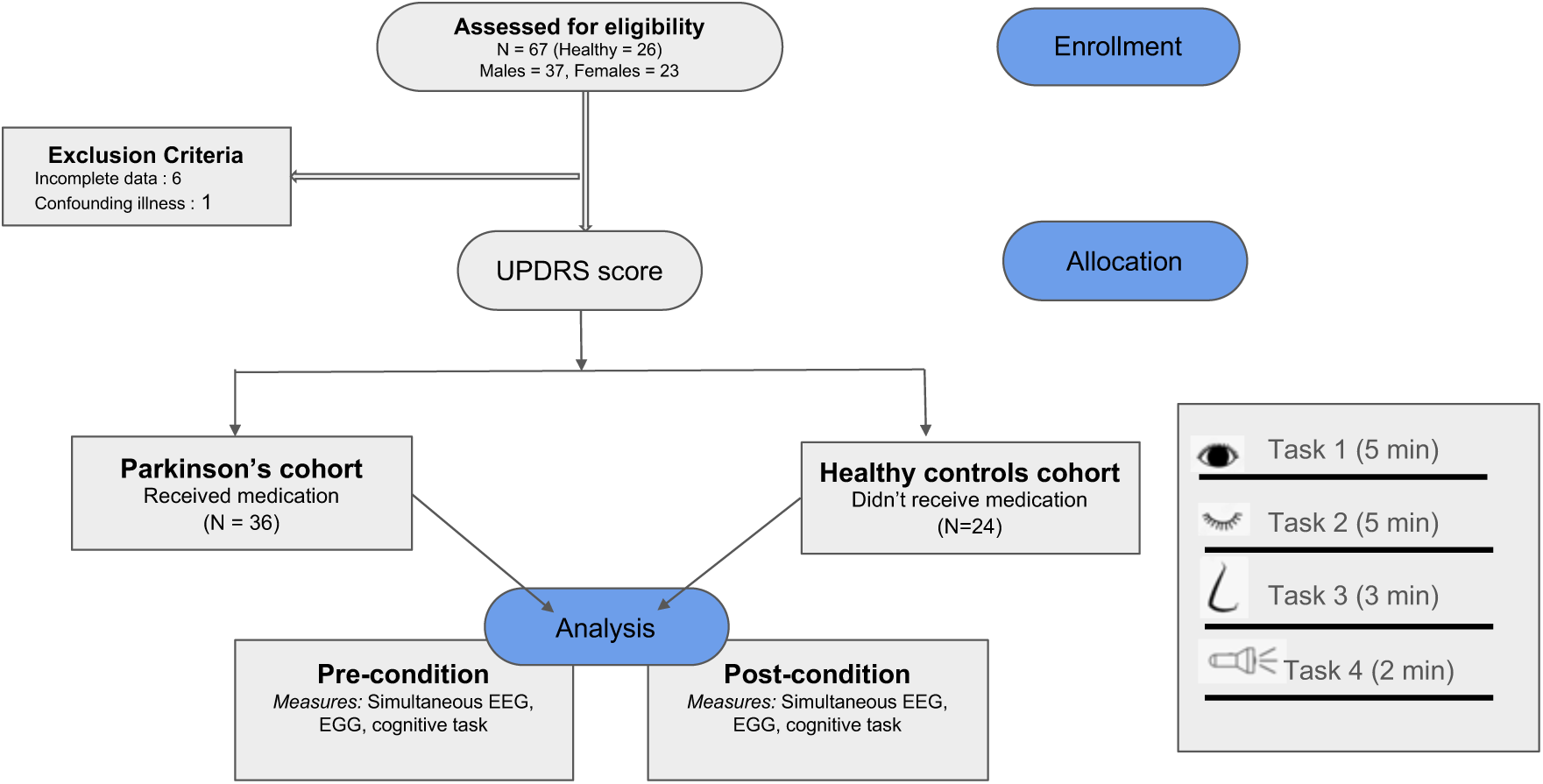
Specifics of our study. a) CONSORT chart b) Study design representing the cognitive tasks conducted during pre- and post-intervention. In Task 1, the participants are asked to be in Eyes open resting state, Task 2 in Eyes closed resting state, Task 3 in hyperventilating state, Task 4 administers photic flickering stimulation at various frequencies. Both pre- and post-intervention were recorded post-prandially at least two hours after meal, and the interventions broadly represent before and after medicine in case of PD, before and after light snacks in case of controls.

**Figure 2:**
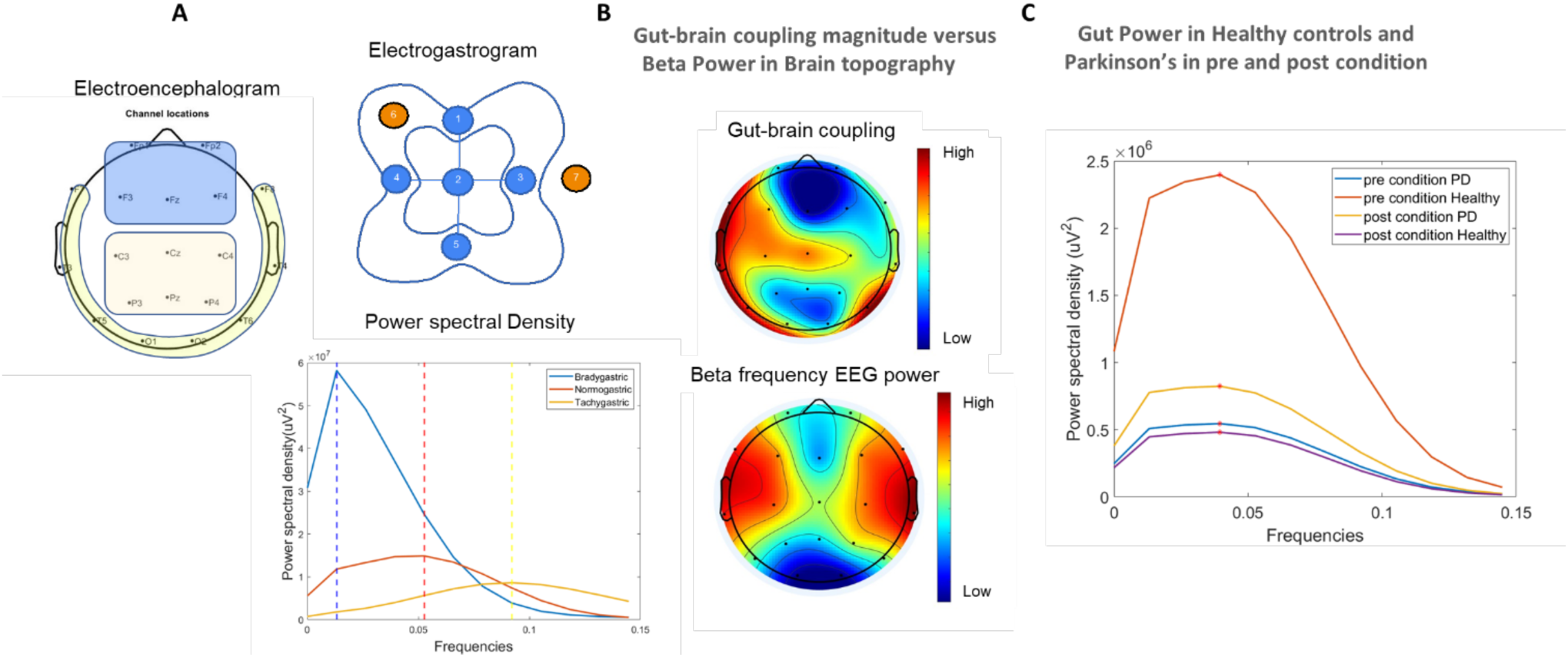
Data acquisition in EEG and EGG. a) Presents a schematic of the recording setup showing the EEG and EGG. Electrode positions, regional grouping into frontal, centro-parietal, and temporo-occipital in the EEG are presented. In EGG channels 2-4 were active channels, with channel 7 as reference, channel 6 as ground. Channel 1 was read in a bipolar reference mode, referenced further to active channel 5. Also shown are the population power spectral density of EGG signals filtered in bradygastric, normogastric, and tachygastric frequency bands. b) shows the phase amplitude coupling between the broadband EGG gut signals and the beta EEG power in various brain regions, and compares it to the channel beta power distribution in EEG. c) shows the population normogastric frequency power in pre- and post-intervention settings in PD and controls, with significant difference in delta intervention (post-pre) between the cohorts (Table 1), all measures were made in post-prandial settings.

**Table 1:**
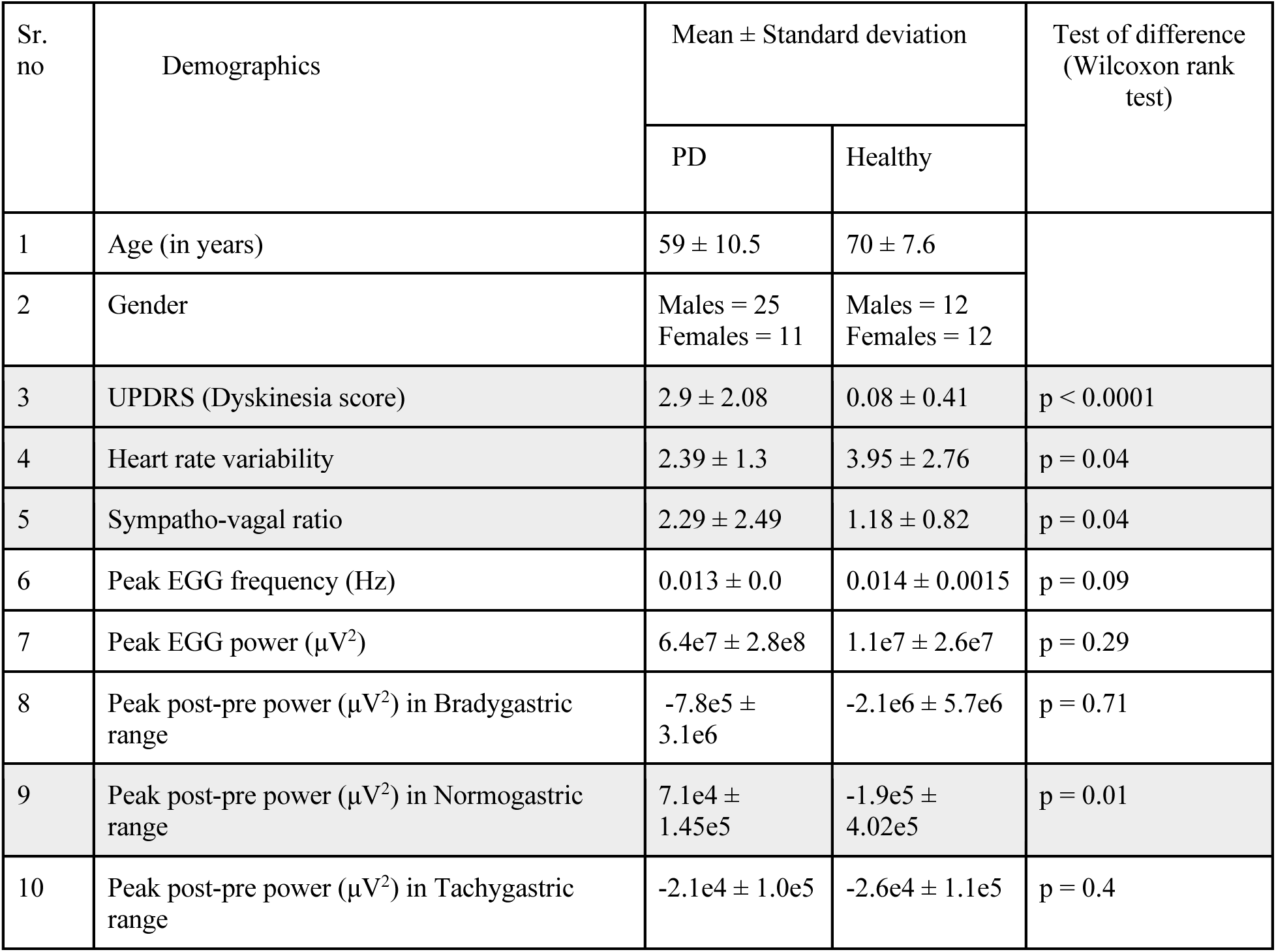
Demographics and basic physiology differences. Rank test statistics suggest the presence of significant differences between PD and healthy controls in terms of the dyskinesia score (UPDRS). The heart rate variability, sympatho-vagal ratio and the medication induced changes in Normogastric EGG Power were also observed to be important differentiating factors.

Our initial analysis further show a robust separation at group level between cohorts on dyskinesia scores (sum of questions 4.1 and 4.2 in UPDRS (Movement Disorder Society Task Force on Rating Scales for Parkinson’s Disease, 2003), heart rate variability, sympatho-vagal balance representing low frequency:high frequency ratio of the Heart rate variability (LF-HF ratio, Table 1). Furthermore in dyskinesia group, we computed the log odds for the forward and backward propagating travelling wave in various gut frequency bands, by stacking signals from EGG electrodes 1, 2 and 4 (forward direction) and performing 2D-fast Fourier transform for understanding the spatio-temporal oscillations (Alamia & VanRullen, 2019). We found that the odds for forward traveling wave propagation in dyskinesia were lesser than the non-dyskinetic groups after intervention (Post-Pre intervention in Tachygastric band, log odds in dyskinesia: −0.0378 ± 0.1356, non-dyskinetic: 0.0123± 0.1062, p=0.07, ranksum test).

### Pre versus post intervention differences were selectively seen in dyskinesia patients

We asked whether the gut-brain coupling strength can significantly explain dyskinesia severity in our participants (N = 31 dyskinesia, 29 non-dyskinesia). We address the question by conducting a repeated measures analysis of variance (rmANOVA) model to explain the coupling strength using cohort status (dyskinesia versus non-dyskinetic cohort), cognitive task identifier, pre or post intervention identifier (referred just as *intervention* here), EGG frequency band identifier, EEG brain region identifier, and their interactions. The model showed main effects of the cognitive task type (F(3,150)=94.33, p<0.00001, η^2^=0.83), EGG frequency band (F(2,100)=234.17, p<0.00001, η^2^=0.89) and brain regions (F(2,100)=13.68, p<0.00001, η^2^=0.32). We also found significant interaction effect between the intervention and dyskinesia cohort status (F(1,50)=5.85, p=0.019, η^2^=0.09). By performing the posthoc Tukey Kramer test, we found that the cohort with dyskinesia had a significant interventional difference (p=0.004). There was also a significant interaction between cognitive task and intervention (F(3,150)=4.02, p=0.01, η^2^=0.18). Posthoc tests revealed that it was primarily the hyperventilating task that showed a significant pre and post interventional difference in dyskinesia group (p=0.001). Similar results were obtained when the cohort status was set to differentiate Parkinson’s patients from healthy controls (refer Supplementary material A).

For the EEG power on the other hand, we found significant main effects of the dyskinesia cohort status (F(1,56)=10.54, p=0.002, η^2^=0.15), intervention (F(1,56)=8.84, p=0.004, η^2^=0.13) and cognitive task type (F(3,156)=6.28, p=0.0005, η^2^=0.25). We also found a significant interaction effect between the cognitive tasks and the brain regions (F(6,336)=5.21, p=0.0004, η^2^=0.37). The posthoc Tukey Kramer test revealed that there was a significant difference in power for the resting state task with closed eyes between centro-parietal and temporo-occipital brain regions (p=0.03).

Interestingly, the normalized EGG power also showed main effects for the cognitive task type (F(3,153)=4.97, p=0.003, η^2^=0.21) and gastric frequency range (F(2,102)=2261.3, p<0.00001, η^2^=0.99). We found significant interaction effects in the frequency bands with the intervention condition (F(2,102)=4.54, p=0.018, η^2^=0.14). Posthoc tests confirmed that there were significant interventional effect for the normogastric EGG power (p=0.002). There was also a significant interaction effect between the frequency bands and the cognitive tasks (F(6,306)=4.96, p=0.0008, η^2^=0.36). Posthoc tests showed that there were significant (p<0.05) differences between photic stimulation and both eyes open, and interoceptive breathing tasks in the normogastric and tachygastric EGG power.

### Normogastric-Frontal coupling could predict dyskinesia severity during interoception

We next asked whether the coupling with the brain could predict the severity of dyskinesia in patients in a continuous manner, for which we developed a robust linear regression model. The model uses coupling strength in the pre-intervention, and the post-pre intervention difference (delta-condition) in subjects to predict dyskinesia severity. A forward sequential feature selection algorithm was used to select the model features using AIC criterion (Table 2).

**Table 2:** Features sensitive to the severity of dyskinesia scores. Significant gut-brain coupling in normogastric ranges were able to predict dyskinesia severity using a robust linear regression model. The coefficients and statistics of the model are provided. The first part of the name denotes the EGG range followed by the brain region. Delta condition refers to the difference due to intervention (post-pre intervention). The cognitive tasks are as follows : T1: Eyes Open, T2: Eyes Closed, T3: Hyperventilating, T4: Photic stimulation.

| Sr. no. | Gut-brain coupling feature | Coefficient | Standard error | p-value |
| --- | --- | --- | --- | --- |
| 1 | Tachygastric-TemporoOccipital (delta condition, T4) | 1.56 | 0.9 | 0.084 |
| 2 | Normogastric-Frontal (delta condition, T3) | 1.29 | 0.59 | 0.029 |
| 3 | Normogastric-Frontal (pre-intervention, T2) | -1.23 | 0.22 | <0.0001 |
| 4 | Bradygastric-Frontal (pre-intervention, T4) | 0.64 | 0.45 | 0.481 |
| 5 | Bradygastric-CentroParietal (pre-intervention, T4) | 0.37 | 0.52 | 0.158 |

The regression model had an adjusted R^2^ value of 0.38 and a RMSE value of 0.19. The significant predictors of dyskinesia symptoms were the change in coupling in the normogastric range of EGG to the frontal brain region especially in the eyes closed (β = −1.23, p<0.001, f^2^ =1.1) and hyperventilating states (β = 1.29, p = 0.029, f^2^ = 0.14). A spider plot of these features for different categories of our population are represented in (Figure 3a, b, e).

**Figure 3:**
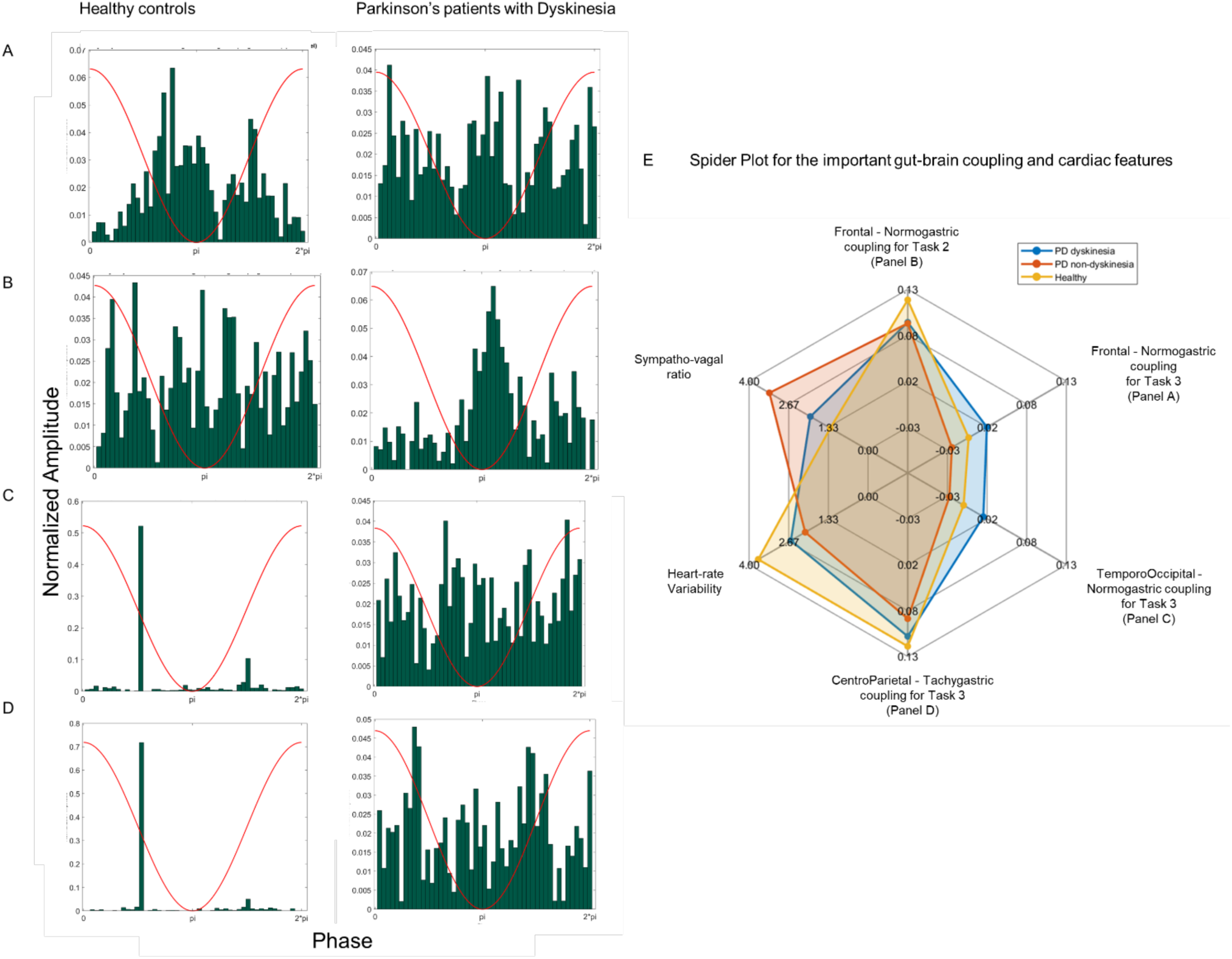
Gut-brain coupling can differentiate PD dyskinesia from rest. The figure presents various significant features identified through regression models. The histogram of EEG amplitude on the EGG phase for a) Normogastric coupling with frontal brain during hyperventilation state (Healthy subject participant id: 1038; LID patient id: 1019) and (b) eyes closed resting state (Healthy subject participant id: 1052; LID patient id: 1012). c) normogastric coupling with temporo-occipital brain during hyperventilation (Healthy subject participant id: 1061; LID patient id: 1026), d) tachygastric coupling with centro-parietal brain during hyperventilation in the pre-intervention (Illustrative Healthy subject participant id : 1052; LID patient id : 1012), shows characteristic coupling differences between dyskinesia and non-dyskinesia groups. e) A spider plot of various gut-brain coupling measures and cardiac features selected from our models that differentiating dyskinesia, non-dyskinesia PD, from healthy controls. The task 2 represents eyes closed resting state, and task 3 is the hyperventilating paradigm. We observed a decreased heart rate variability and an increased sympatho-vagal ratio in PD compared to controls.

Lastly, we added demographics informative of age and gender and cardiac features as covariates to understand their explanatory effects. We did not find any age, gender, HRV, sympatho-vagal ratio interactions with these key features.

### Coupling with centro-parietal and temporo-occipital brain regions assists in differentiating dyskinesia condition

Can gut-brain coupling measures classify people with dyskinesia symptoms from healthy controls and non-dyskinetic PD at a broader level? We answered this question using a logistic regression model, and find that the coupling between gut and brain in various contexts were indeed able to predict the presence or absence of dyskinesia in our population.

The model features include the measures computed in the pre-intervention, and the post-pre intervention (delta-condition) observed in subjects, and the accuracy scores were calculated by repeating stratified 5-fold cross validation 20 times. The model had a F1 score of 0.7, MCC score of 0.46, specificity and sensitivity of 0.78 and 0.67 respectively.

The critical features predicting the dyskinesia group from the non-dyskinesia group include the normogastric coupling with the frontal and the temporo-occipital cortices, and the tachygastric coupling with the centroparietal region (Table 3, Figure 3c,d). The first two covariates were significant especially in the eyes closed (β =−12.3, p =0.046, f^2^ =0.08) and hyperventilating cognitive state (β =26.3, p = 0.003, f^2^ =0.22) respectively, while the centroparietal feature were seen in the hyperventilating cognition (β =−13.7, p = 0.005, f^2^ =0.17). There were no significant interactions with age, gender.

**Table 3:** Features differentiating the non-dyskinetics from dyskinesia patients. Significant gut-brain coupling in normogastric and Tachygastric ranges were able to broadly predict dyskinesia population from non-dyskinetic population using a robust logistic regression model. The coefficients and statistics of the model are provided. The first part of the name denotes the EGG range followed by the brain region. Delta condition refers to the difference due to intervention (post-pre intervention). The cognitive tasks are as follows : T1: Eyes Open, T2: Eyes Closed, T3: Hyperventilating, T4: Photic stimulation.

| Sr. no. | Gut-brain coupling Feature | Coefficient | Standard error | p-value |
| --- | --- | --- | --- | --- |
| 1 | Normogastric-TemporoOccipital (delta-condition, T3) | 26.3 | 8.9 | 0.003 |
| 2 | Tachygastric-CentroParietal (pre-intervention, T3) | -13.7 | 6.7 | 0.005 |
| 3 | Normogastric-Frontal (pre-intervention, T2) | -12.3 | 6.2 | 0.046 |
| 4 | Normogastric-TemporoOccipital (pre-intervention, T2) | 4.0 | 6.7 | 0.554 |
| 5 | Normogastric-Frontal (pre-intervention, T4) | -1.2 | 4.0 | 0.763 |

While larger coupling marks PD severity in the tachygastric range during hyperventilating state, less coupling relates to PD severity in the temporo-occipital area-normogastric connection during the hyperventilating state. Moreover, normogastric coupling with frontal region are sensitive to predicting continuous dyskinesia scores for PD.

### Is the brain a mediator for levodopa induced effects from gut?

Finally we asked a more specific question—whether the gut has a directional effect on dyskinesia with the brain as a mediator? The implication of the answer could be profound to answer for targeted interventions. Moreover from the earlier analysis, we find that the distinct task settings differentially predict the dyskinesia scores. So, we conducted multilevel mediation analysis (see Methods) to understand the effect of brain EEG power as a mediator between EGG power and dyskinesia severity in the PD population, with tasks as the second level used for mediation.

Across the tasks in the second level, the relationship between tachygastric gut power and Dyskinesia score is mediated by centro-parietal beta power (a =−0.16±0.06, p=0.001; b= −1.05±0.2, p<0.0001; ab=−0.11±0.03, p=0.001, cross-validated using permutation test). All of these signals were taken at the pre-intervention and in dyskinesia cohort (Figure 4).

**Figure 4:**
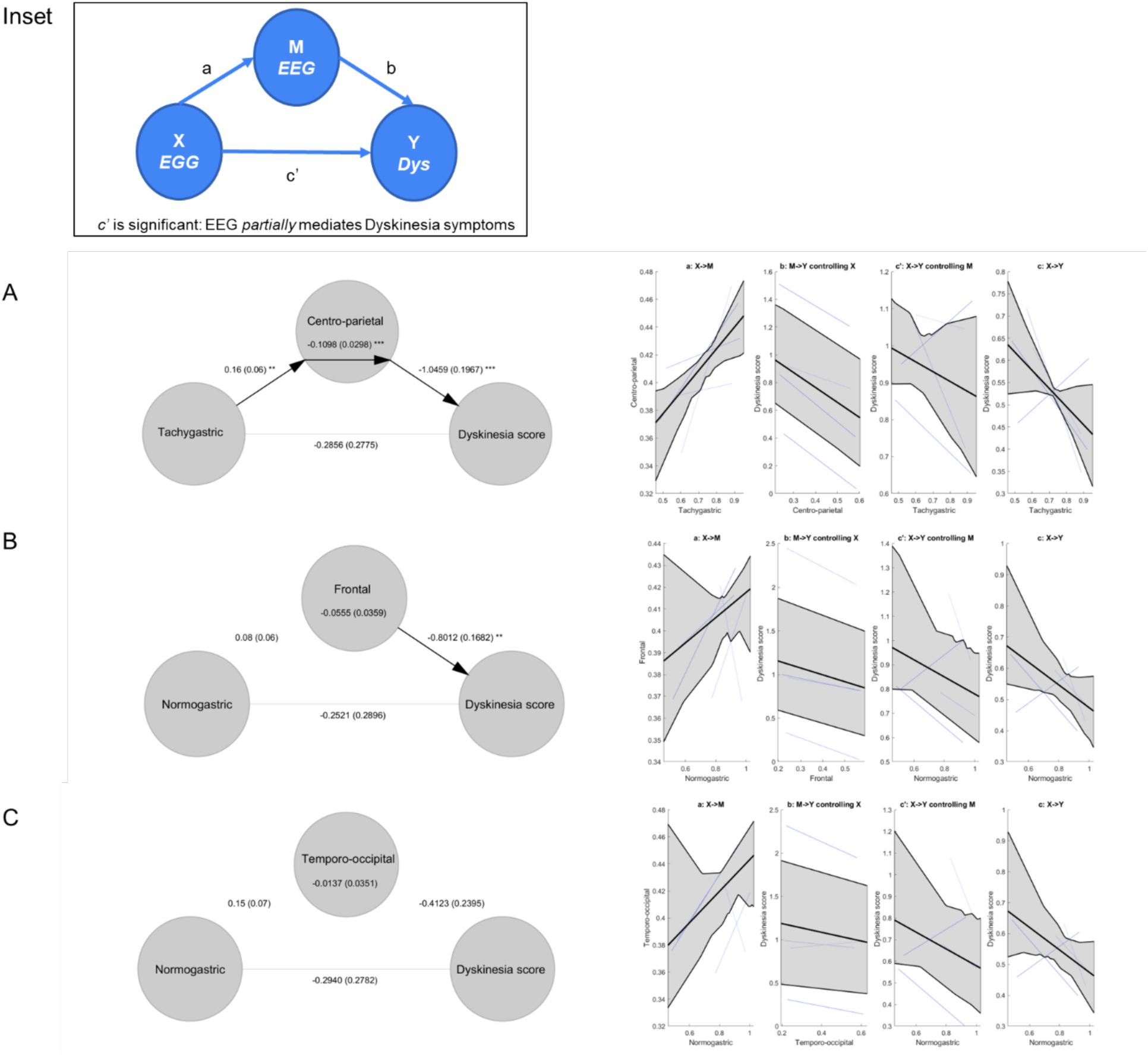
Mediation analysis. The inset describes the mediation model structure relating the power in gut, brain, and severity of dyskinesia. If a and b are significant, and c’ is insignificant, the gut activity partially mediates the dyskinesia symptoms by interacting with the brain. (a) We observe the complete mediation of tachygastric gut activity interaction with centro-parietal brain to explain dyskinesia severity. (b, c) Presents the mediation results for normogastric gut activity interaction with frontal activity and with the temporo-occipital activity, respectively. The right side panels provide evidence of various directional relationships between X (gut activity power), M (brain activity power), and Y (symptom severity for dyskinesia). The black lines represent the mean values of coefficients, the grey shaded areas represent the 95%ile of those values and the blue lines represent the coefficient values for the individual cognitive tasks.

### Heart rate variability differs with various Normogastric-Frontal and Tachygastric-CentroParietal coupling levels to explain the dyskinesia symptoms

Heart rate variability (HRV) is a measure of the variation in time intervals between consecutive heartbeats. It reflects the adaptability and flexibility of the cardiovascular system in responding to internal and external stimuli. HRV is influenced by the interplay between the sympathetic nervous system (responsible for the “fight or flight” response) and the parasympathetic nervous system (responsible for the “rest and digest” response) among other physiological factors (Tiwari et al., 2021).

We found significant interactions of baseline heart rate variability (HRV) with the gut-brain coupling features broadly distinguishing dyskinetic group from of the rest in the logistic regression model. Marginal effects were computed to precisely probe the interactions: The effect of pre-intervention HRV on the predicted probability of presence of dyskinesia was derived at different levels of the coupling feature while keeping all the other features constant at their mean values. We also computed the otherwise: the effect of gut-brain coupling on the predicted probability of dyskinesia presence at different levels of pre-intervention HRV (Figure 5). For the four critical features identified from logistic and linear models described earlier for predicting dyskinesia, we found that the interaction of baseline heart rate variability (HRV) with the normogastric coupling to the frontal cortical region (β =190.7, p = 0.013, f^2^ =0.19) and the tachygastric coupling to the centro-parietal cortical region (β =−149.6, p = 0.018, f^2^ =0.18) in the hyperventilating task were significant.

**Figure 5:**
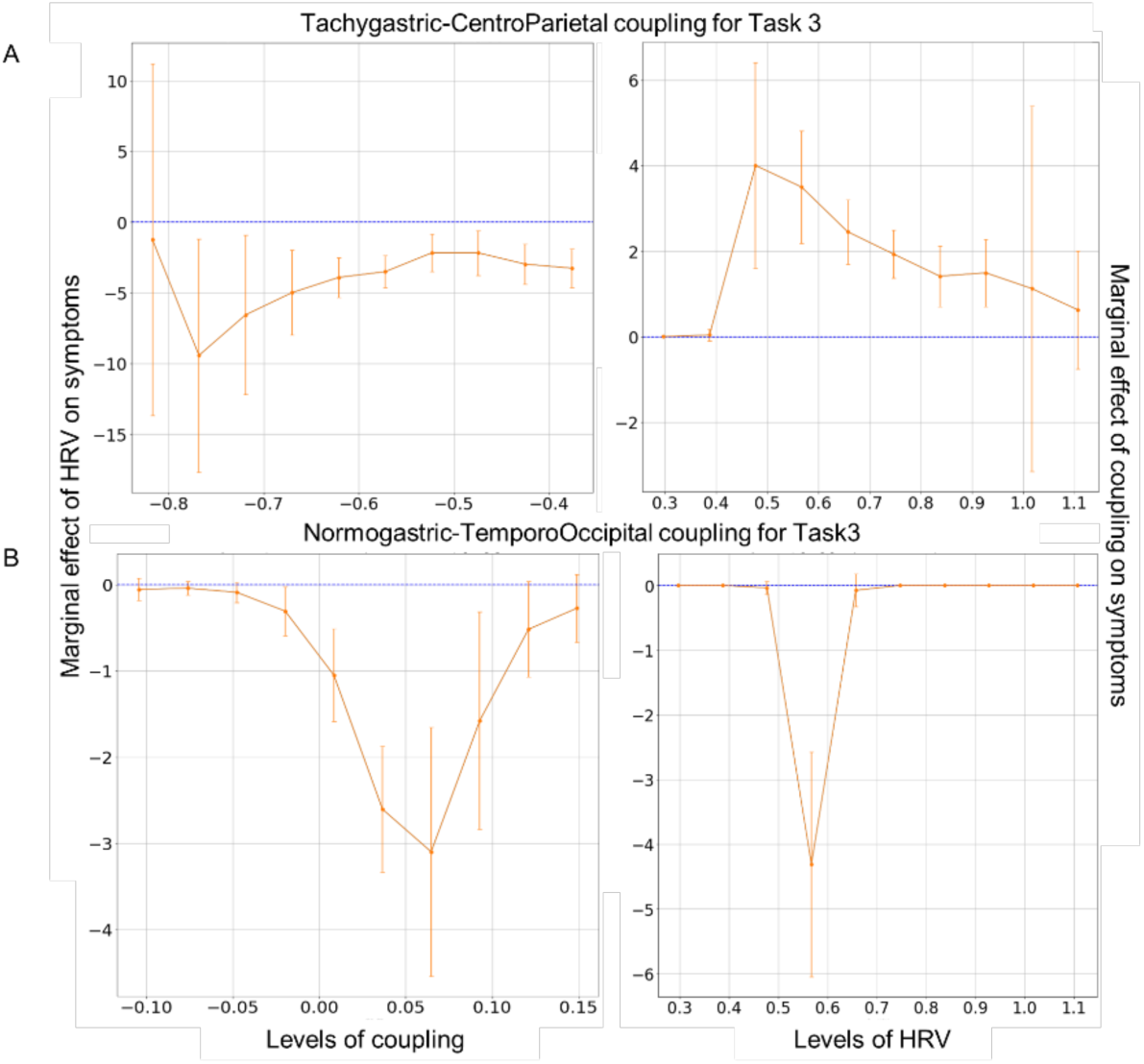
Marginal effects of HRV in understanding dyskinesia. Presents the effect of varying levels of HRV to understanding the effect of gut-brain coupling on dyskinesia in the left, and the effect of varying levels of coupling to understanding the effect of HRV on dyskinesia on right. (a) shows the results for tachygastric coupling to the centro-parietal cortical region. (b) the results for normogastric coupling to the frontal cortical region. The model can be used to guide interventional tuning of HRV to arrive at the sensitive ranges estimated to modulate the severity of symptoms.

## Discussion

The study has two main outcomes, first: electrogastrogram (EGG) as a technique can be used to study gut activity and gut-brain coupling when simultaneously recorded with electroencephalography (EEG), and second: the method is sensitive to Parkinson’s disease severity and relates to levodopa induced dyskinesia. This is in line with some recent observations by Araki et al, who showed that EGG could be used as a potential tool for PD diagnosis (Araki et al., 2021), and can be sensitive to changes in gut rhythms (Naftali et al., 2005).

All the participants were studied post-prandially, with simultaneous EEG and EGG data collected during various cognitive tasks in *pre-intervention condition* and after a perturbation in *post-intervention condition* with intake of medication in case of patients and some light snacks in case of healthy controls. In our population sample residing in Northern part of India, we observed that the average peak EGG frequency is in the bradygastric range for both PD and healthy cohorts. Interestingly this slowing of gut was earlier shown by recent local surveys in the country that reported a whooping ∼60% people having gut motility issues at least once a week (Chen et al., 2005; Ghoshal et al., 2018; Rajasekhar, n.d.; Zou et al., 2024). Upon intervention, our study finds significant changes (reported as delta condition) in PD, with post-interventional power larger than the pre-intervention.

We compute gut-brain coupling using phase amplitude coupling method, where the strength indicates the extent to which phase of the gut signal is related to the amplitude of the brain signal (Özkurt & Schnitzler, 2011). We used the coupling strength to investigate whether they explain the severity of dyskinesia symptoms individually, in three different gut frequency bands— bradygastric (slow gut oscillation activity), normogastric (normal gut oscillations), and tachygastric (fast gut oscillations). The analysis was limited to the beta frequency of the brain signal, with the brain categorized into 3 major regions such as frontal, centro-parietal, temporo-occipital, for analysis simplicity.

We invoke various cognitive paradigms, inspired by the earlier literature, to control the gut-brain coupling. Some include being in resting state in eyes open fashion, in interoceptive states where we broadly include being in eyes closed resting state in a quiet place or those invoking attention to breathing in hyperventilation state (Ritz et al., 2024), and lastly a visual flicker paradigm (Bartley, 1939). Evolutionarily, the occipital areas have been related to gut activity for acting as an important sensory module to associate with navigation planning and foraging behavior (Azzalini et al., 2019; Rebollo et al., 2018, 2021). Not surprisingly for this reason, Temporo-occipital was one sensitive brain region to explaining the gut-brain coupling in our study. Relatedly, our study finds that the hyperventilating paradigm and the eyes closed resting states were more sensitive to dyskinesia measurements, suggesting the significance of interoception in PD evaluation and prognosis.

We find all three major brain areas used in our study to be related with dyskinesia. Centro-parietal region is one significant area housing the motor strip, that is believed to be mediating the motor complication (Donoghue & Sanes, 1994; Rizzolatti & Luppino, 2001). The involvement of frontal cortices in dyskinesia has been noted in many earlier studies as well, that mostly proposes the involvement of inferior frontal gyrus and the basal ganglia circuity (Barroso-Chinea & Bezard, 2010; Cerasa et al., 2015; Hulme et al., 2022; Petersson et al., 2020). Our study also suggests that normogastric gut coupling to frontal beta activity in the brain can reliably predict dyskinesia symptom severity in Parkinson’s Disease.

More interestingly, we observe the effect of HRV can modulate the gut-brain coupling and thereby the dyskinesia severity using a mediation analysis. We also computed the marginal contribution of different coupling features to dyskinesia severity at different levels of heart rate variability. We observe some sensitive ranges of HRV for the gut-brain coupling to effectively reduce dyskinesia. We propose these models are a guiding tool to optimize the setup of various intervention methods altering cardiac signals including the vagal nerve stimulation for improving the dyskinesia symptoms (Sigurdsson et al., 2021). Further studies are needed to validate the model’s utility for intervention.

The major limitations of our study include small population size with low number of non-dyskinetic PD patients, and having broader regions of investigation of the brain signals for analysis simplicity. Future work may also look at specific brain sources and detailed gut motility patterns for precise modeling. We did not see any order effect in our data, however, future work could also randomize the order of cognitive tasks with larger sample size for increased robustness in the study observations. Overall, our results show that frequency specific electrogastrography during interoception could be a potential way to predict the dyskinesia severity in PD and can be of interventional significance.

## Methods

### Participants

The study involves experiments with two independent groups, one suffering from Parkinson’s that are referred to us by our collaborating neurologist in the city (N = 39, 59 ± 10.5 years) and another healthy control group (N = 28, 70 ± 7.6 years). A clinical diagnosis of idiopathic PD was based on the movement disorders society clinical diagnostic criteria for PD. The patients were prescribed regular anti-parkinsonian drugs, such as Syndopa and Parkinta or Xafinact and dopamine agonists along with some vitamins. The clinical stage of the patients was assessed using United Parkinson’s Disease Rating Scale motor score (UPDRS). The control subjects were those who had not been diagnosed with PD. We also excluded participants who could not cooperate or had some other neurological illness including epileptic-type seizures.

### Sample size

We targeted a sample size of N=70 (Power 0.8, α = 0.05, repeated measurements ANOVA with pre and post gut intervention, assuming moderate effect size f=0.25 and correlation about 0.5) for referral from the collaborating neurologist, and for healthy control recruitment. We managed to recruit a total of 67 participants over a period of about 10 months, 2 were excluded from further analysis because of missing data files and 1 due to post-hoc observation of not satisfying exclusion criteria. Four more participants were later excluded due to missing EGG simultaneous recording data. The final cohort consisted of 60 subjects (N=36 with Parkinson’s disease, N=24 Healthy controls).

### Ethics approval

The experiment was designed and conducted in compliance with protocols approved by the Institute Human Ethics Committee of Indian Institute of Technology, Kanpur. Also, a prior written informed consent was taken from the participants.

### Experimental Design

The experimentation was performed in a pre-post design, with electroencephalography (EEG) and electrogastrography (EGG) collected two times (*pre and post intervention*). The participants walked into the clinic after they had breakfast at least before 2 hours, and were in the post-prandial state. The pre and post intervention were separated by about 20-30 mins duration, during which medication for PD patients, and light snacks for the healthy controls were provided. Most of our analysis were done in the data collected during pre-intervention and the post-pre intervention difference (also referred to as *delta-condition*) in our study.

### Cognitive Tasks

The Electroencephalogram (EEG) and Electrogastrogram (EGG) were setup at the beginning of the experiment, and they were maintained in position throughout the experimentation.

We administered the below four cognitive tasks in each of pre and post time period while simultaneous EEG-EGG data was recorded. The first being eyes open resting state (T1 for 5 mins) at an experimental room in the clinic of our collaborating neurologist. Next was being in the eyes closed resting state (T2, 5 mins), third was asking the participants to hyperventilate by letting them breathe deep and holding it for about 5 secs for *ten* times (T3, 2:30 - 3mins). Finally, we administered photic stimulation of 3,5,7,9,11,13,15,17,19,21 Hz for 10 secs each and inter trial interval of about 1 sec between frequencies (T4, 2 mins), by locating the machine (photic emitter) at one side of the subject at about 50 cm distance at an altitude matching the subject’s face location.

### Electroencephalography acquisition

We used a 19 channel, portable, low power recording setup from Clarity medicals of India (BrainTech 24+) for acquiring EEG signals from our participants.

The specific preprocessing steps are: the EEG data was downsampled from 256 Hz to 250 Hz. Three seconds of data at the end was removed since they were recording artifacts (flat line). The data was bandpass filtered between 1-45 Hz (the *pop_eegfiltnew* command in EEGLAB was used with the *filtorder* parameter set to 9000). Channel data which record noise for extended periods of time were removed using the *clean_channels* function of EEGLAB (Delorme & Makeig, 2004). Next, Independent component analysis (ICA) was done on the remaining data. The *ICLabel* function (Pion-Tonachini et al., 2019) was used to calculate the confidence percentage of different sources viz., the brain, muscle, heart, ocular, line noise, channel noise and other. If any of the components had a confidence of percentage for the brain source less than 5%, they were removed. We used the *clean_rawdata* function to remove portions of the data that had non-repeating and non-stationary artifacts, which couldn’t be captured by ICA, using the Artifact Subspace Reconstruction (ASR) algorithm. The algorithm first identified a clean stretch of data (calibration data) and rejected any portion of the data if the standard deviation exceeded 20 times the standard deviation of the calibration data (9.3±1.6 % of the data). The channel data which were removed (2.5±0.9 channels) at an earlier stage of the preprocessing were interpolated using the *pop_interp* function which used spherical spline interpolation for the same. The data was then average referenced and stored as .set file for further analysis.

We used Welch’s periodogram method of time frequency decomposition (using *pwelch* command in MATLAB, window size of 5 seconds with 50% overlap) to get the power spectrum for the EEG signal, which was then normalized by dividing by the sum of the entire spectrum. The relative power in the beta frequency band (13-30 Hz) was calculated by dividing the sum of power in those frequency bands by the total power in the entire 1-30 Hz spectrum. We chose beta band frequency for its significance in representing motor functions (Muralidharan & Aron, 2021a, 2021b; Picazio et al., 2014).

We primarily focused on 3 broader regions of the cortex (ROIs) for its coupling with the gut (Rebollo et al., 2018), such as 1) Frontal (mean of channels FP1, FP2, F3, F4, Fz), 2) Temporo-occipital (F7, T3, T5, O1, O2, T6, T4, F8), and 3) Centro-parietal (C3, Cz, C4, P3, Pz, P4).

### Electrogastrography acquisition

The electrogastrogram (EGG) was acquired with 7 disposable cutaneous electrodes (OpenBCI make, 5 active, 1 reference, 1 ground) situated on the abdomen and recorded simultaneously with the EEG. The channel 1 was placed about 5 cms below the Xiphoid process of the sternum, and the others were separated by radius of approximately 5cms as shown in *(Figure 1b)*. Data from channels 1-5 were referenced using channel 7, channel 6 was used as ground, and channel 1 was recorded in a bipolar mode (referenced to channel 5).

The raw EGG signals were filtered in the three ranges: bradygastric (0.0083-0.03 Hz), normogastric (0.03-0.07 Hz) and tachygastric (0.07-0.15 Hz), cutting down the respiratory (0.2-0.4 Hz) and cardiac (1-1.7 Hz) artifacts (Wolpert et al., 2020).The MATLAB command *bandpass* was used to filter the EGG signals. This designs a minimum order bandpass FIR filter with the specified upper and lower frequencies. Power spectral density (*pwelch* command in MATLAB was used with an one minute window and 50% overlap) was calculated for all the EGG electrodes and it was normalized by the sum of the entire spectrum. The relative power was calculated for each of the EGG ranges by adding the power bins for that spectrum which was divided by the sum of the normalized power spectrum. The signal with the highest relative power across channels was further chosen for the calculation of PAC, for any condition. The time domain signal of this electrode was z-scored.

### Phase amplitude coupling (PAC)

A direct PAC estimate (Özkurt & Schnitzler, 2011) was computed using the following formula:

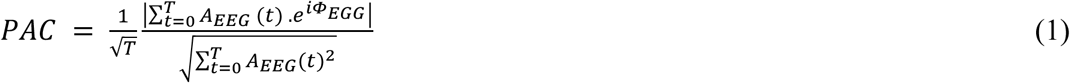

where *Φ_EGG_*(*t*) is the instantaneous phase of the low-frequency oscillation computed in MATLAB using angle() function of the hilbert transformed EGG best-electrode data, and *A_EEG_*(*t*) is the instantaneous amplitude of the high frequency oscillation computed using abs() function of the hilbert transformed EEG data. PAC values were calculated for the 19 channels and then the values corresponding to the channels in the regions were averaged.

### Mediation analysis

In this analysis, we studied whether X had a partial or complete effect on Y, through a mediator, M. This was done by setting up 3 regression equations. Here *i* and *e* were intercepts and errors respectively.

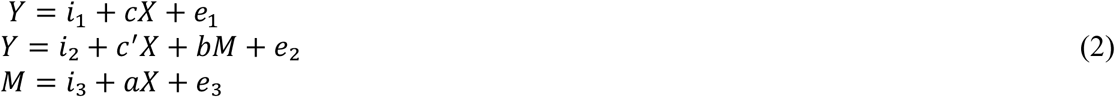

For a mediation to be claimed, *a*, *b* and the product of them *ab* needed to be statistically significant. If *c’* was statistically insignificant, it was called *complete mediation*. If *c’* was significant, then it was called *partial mediation* (Iacobucci, 2008; MacKinnon, 2012).

For our analysis we specifically worked with the *dyskinesia* aspect of PD, and asked whether there were task specific mediations between gut and dyskinesia via the brain activations. We considered a multilevel model with the subjects as a lower level and the four cognitive measurements used as the upper level. All the effects may vary by the second level (by task) and subscripted as *j* in the upcoming equations. Hence the equations that described this multilevel mediation model were as follows:

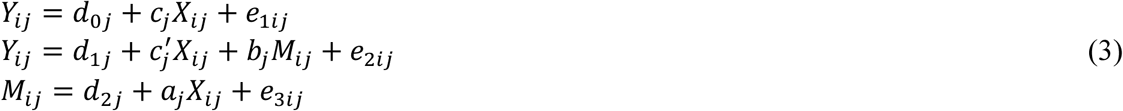

The indirect effect (*a_j_b*_j_) in a multilevel mediation model is *ab* + σ_ab_ where σ_ab_ is the population covariance of *a_j_* and *b_j_*.

In our study, we conducted mediation analysis to look into whether EEG power mediates the effect of gut power on dyskinesia severity, so we assumed X ∼ func(gut power), Y ∼ func(UPDRS score), M ∼ func(EEG power).

### Heart rate variability (HRV) and sympatho-vagal ratio (LF-HF ratio)

Two components of HRV that are often analysed are (Pham et al., 2021) a low Frequency (LF)-LF power within the frequency range of 0.04 to 0.15 Hz, thought to reflect more of a sympathetic modulation. The other one is a high Frequency (HF) component-HF power which majorly reflects parasympathetic modulation of heart rate occurring within the frequency range of 0.15 to 0.4 Hz. The ratio of LF/HF power represents the sympatho-vagal activity ratio.

We extracted the necessary HRV measures from the channel closer to the sternum (*refer to channel 2 in Figure 1b*) with clear RR peaks (refer to Supplementary Material B). We used HRVTool package (Vollmer, 2015, 2019) to analyze the data as waveform at a sampling rate of 250 Hz, for time length of each of our tasks in pre and post intervention, to compute the HRV and the vagal tone measures.

Using the HRV tool, we initially filtered the signals with successive R-R intervals within an interval of *±20 msecs*, referred in the tool as pNNi. The relative R-R intervals of the signals (rr*_i_*) are used to compute the HRV.

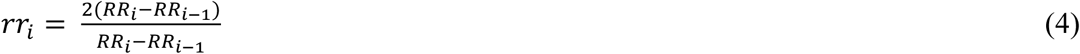

where, n = number of RR intervals, rr describes the relative variation of consecutive RR intervals with distance one, which is usually between −20% and +20%.

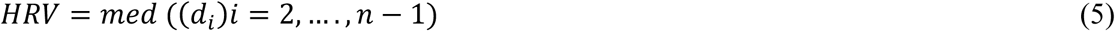

with di as Euclidean distance between (rri; rri+1) and centre point, which is the average of relative RR intervals for which |rr| < 20%.

The spline interpolated R-R intervals in dependence (RR Tachogram) to time are used to compute the power spectrum, specifically in the LF and HF ranges, to compute the balance between sympathetic and vagal tone.

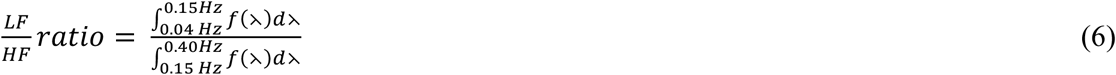

### Travelling wave analysis

We stacked the data from electrodes 1,2,4 arranged in that order on the gut surface, through time, in the pre- and post-intervention. The data was divided into sliding windows of 1 minute without overlap and a 2D Fast Fourier Transform was applied and the log odds for forward and backward travelling wave were calculated for each window according to Alamia et al. (Alamia & VanRullen, 2019). Basically from the resulting 2D spectrogram, the ratio of the mean power in the upper-left and the lower-left quadrants were used to compute the log odds. The log odds for all the sliding windows were averaged to get a single value for each subject.

### Statistical Corrections

The statistical significance of all parameters in the mediation model are determined using bootstrapping method. This is primarily done because the effect *ab* is a non-normal distribution. The bootstrapping was done by resampling 1000 times with replacement of the data at the level of the tasks (2^nd^ level). The alpha value for rejecting the null hypothesis was kept at 0.05. The M3 toolbox in MATLAB was used to perform the multilevel mediational analysis (Wager et al., 2008).

Wilcoxon rank test (signrank test in MATLAB) was used to compare between group Power, gut-brain coupling, HRV and sympatho-vagal features, as the underlying samples were non-normal in distribution. The Cohen’s f^2^ was computed as: (R^2^ of full regression model – R^2^ of reduced model)/(1-R^2^ of full model).

We conducted repeated measures ANOVA using rmANOVA command in MATLAB. For post hoc tests in rmANOVA, we used Tukey Kramer test (executed using *multcompare* command in MATLAB). For two sample hypothesis tests, we used Wilcoxon test (*ranksum* in MATLAB).

### Predictive models

A forward sequential feature selection algorithm (SFA) was used to select the relevant features from the PAC feature set (custom script for sequential feature selection in Python3.8) and predict the dyskinesia specific UPDRS score (sum of sections 4.1 and section 4.2). The pre-intervention features were log transformed and divided by 4 to normalize them in the [-1,1] bounds. The pre-intervention cardiac features (HRV, sympatho-vagal ratio) were considered and they were sqrt transformed and divided by 3 for [-1,1] range normalization. The age was divided by 80 for normalization in the [0,1] range. The dyskinesia score was divided by 8 for the [0,1] range normalization. Missing values were imputed using mean of the feature columns.

*Linear prediction model:* The scoring metric for the linear regression model was a modified version of AIC to account for the small sample sizes (Sugiura, 1978). This was done to maximize the fit to the data while keeping the model parsimonious. Only the PD dyskinesia and non-dyskinesia patients were considered for the linear regression models to improve the model predictability as the healthy people had zero scores.

*Logistic prediction model:* The scoring metric for the logistic regression model (*sklearn* library in Python with a regularization parameter of 1 which indicates a high level of L2 regularization) was 5-fold cross validated accuracy. All subjects were considered and the logistic model was used to differentiate between the subjects with dyskinesia (dyskinesia score > 0) and without dyskinesia (dyskinesia score = 0).

In order to improve consistency of the predictive features for the linear model (logistic model) after accounting for sample bias and random initializations, we performed random selection of about 90% of samples without replacement about 100 times and SFA was applied by minimizing the AIC score with a tolerance of 5 points (cross validated accuracy with a tolerance of 0.03 points). The ones repeating more than ⅓ of the total repeats were taken as the new features for the model.

## Data Availability

Data will be shared upon reasonable requests to the corresponding author.

## Code Availability

The codes used in the manuscript are available in our lab Github link here: https://github.com/PragathiBalasubramani/TransitLab

## Acknowledgements

We sincerely thank Prof. Srinivasa Chakravarthy and Anirban Bandyopadhyay and all the members of our Translational Neuroscience and Technology group for helpful discussions. We also thank those clinical personnel assisted in data collection, and all our participants for kindly agreeing to our study.

## Author information

PPB, NPB conceptualized the study. HPT performed data collection and data management. SH and MK performed the data analysis. SH and PPB wrote the first draft of the manuscript, proofread the document and prepared the final draft.

## Supplementary Material A

### Repeated measures ANOVA model

#### PD v/s Healthy

With a repeated measures ANOVA model for understanding the PD versus healthy group effects, We found main effects of the cognitive tasks (*F(3,150)=90.22, p < 0.00001, η^2^=0.83*), brain regions (*F(2,100)=11.18, p < 0.0001, η^2^=0.28*) and EGG ranges *(F(2,100)=212.46, p <0.00001, η^2^=0.88*). There was a significant interaction effect between the intervention and the cognitive tasks (*F(3,150)=3, p=0.03, η^2^=0.14*). The posthoc Tukey Kramer test revealed that there were post and pre-intervention differences in the hyperventilating breathing task (p=0.007).

In terms of EEG beta power on the other hand, we observed significant effect of cohort status (*F(1,56)=12.23, p=0.0009, η^2^=0.17*), and whether the EEG was conducted in pre or post intervention (*F(1,56)=9.88, p=0.003, η^2^=0.15*). This is in addition to the task effects on EEG (*F(3,168)=7.9, p < 0.0001, η^2^=0.3*). Further, there was a significant interaction between the cohort status and the cognitive tasks (*F(3,168)=2.9, p=0.037, η^2^=0.13*). The posthoc Tukey Kramer test revealed that there were no significant (p>0.05) differences in EEG power of different tasks for the PD cohort while there were significant (p<0.05) task power differences in the healthy controls. There was also a significant interaction between the cognitive tasks and the brain regions (*F(6,336)=5.02, p=0.0005, η^2^=0.36).* When the posthoc Tukey Kramer test was done, we found that the centro-parietal and temporo-occipital brain regions have significant (p<0.05) power differences between the cognitive tasks while the frontal region lacked any such differences.

The EGG power showed differences due to task (*F(3,153)=5.12, p=0.002, η^2^=0.22)*, and the frequency bands (*F(2,102)=2138.6, p<0.00001, η^2^=0.97*). There was a significant interaction effect between cognitive tasks and frequency bands (*F(6,306)=4.14, p=0.003, η^2^=0.32*). The posthoc Tukey Kramer test revealed that there were significant (p<0.05) power differences between tasks in the normogastric range while the tachygastric and bradygastric ranges had no significant (p>0.05) taskwise power differences.

## Supplementary Material B

**Figure S2a:**
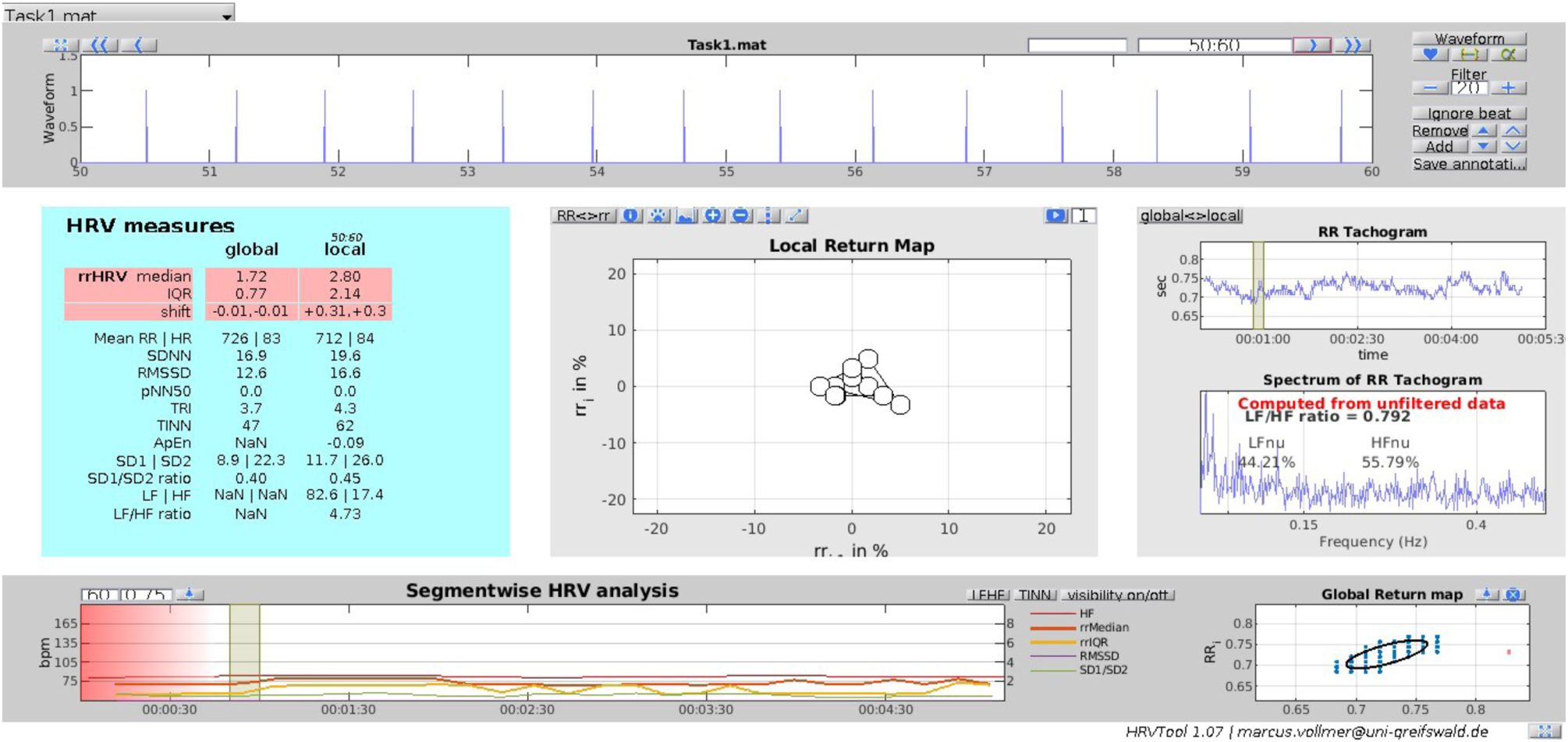
Illustrative PD participant characteristics obtained through a HRV tool (patient id: 1005).

**Figure S2b:**
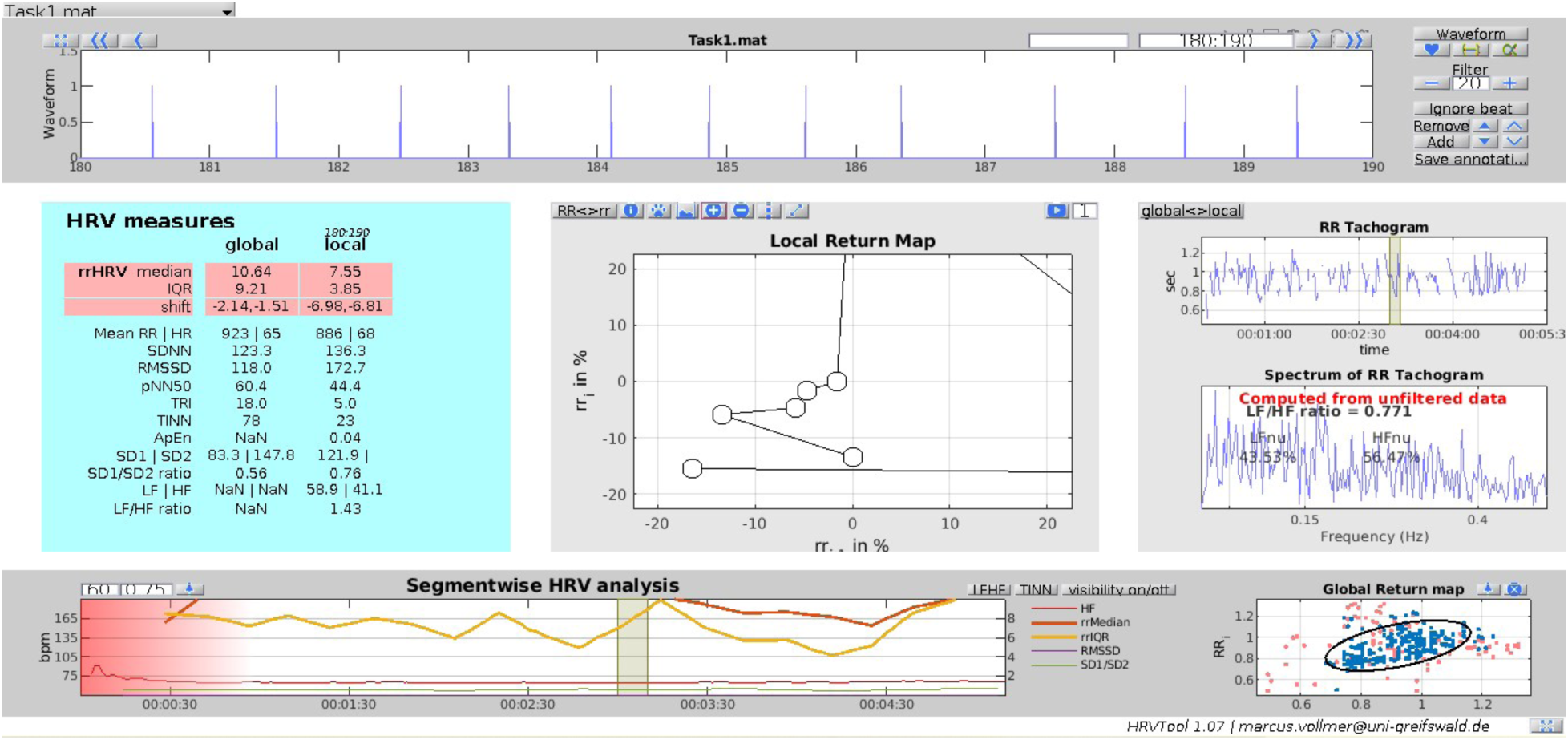
Illustrative healthy participant characteristics obtained through a HRV tool (participant id: 1043).

